# Impact of Breast Cancer on Body Image: A Systematic Review and Meta-Analysis

**DOI:** 10.64898/2026.03.17.26348652

**Authors:** Serenay Yazici Sarikaya, Rui Wang, Ann-Christin S. Kimmig, Sara Brucker, Markus Hahn, Anna Wikman, Birgit Derntl

## Abstract

**Purpose:** Breast cancer is the most common cancer among women worldwide. Although treatment is essential and often life-saving, it can have profound consequences beyond physical health, particularly for psychological well-being and body image. This study aimed to systematically review and synthesize the existing evidence on how breast cancer and its treatment affect body image in women.

**Methods:** We conducted a systematic search of four major electronic databases for peer-reviewed studies published up to 2025. In total, 41 studies met the inclusion criteria for the systematic review, and 24 were included in the meta-analysis. Standardized mean differences (SMDs) with 95% confidence intervals were calculated using a restricted maximum likelihood model. Analyses were performed in JAMOVI, and heterogeneity was assessed using Cochran’s Q and I² statistics. The review was registered in PROSPERO (CRD42024503033).

**Results:** Females with breast cancer reported significantly lower body image scores than healthy controls (ES =-1.02). Furthermore, body image scores declined from pre-treatment to during treatment (ES =-0.45). Mastectomy was associated with poorer body image compared to breast-conserving therapy (ES =-1-11).

**Conclusion:** Although medically essential, breast cancer treatment can adversely affect body image. Integrating body image support into treatment plans is crucial for promoting the overall health and quality of life of females diagnosed with breast cancer.

## 1. BACKGROUND

Breast cancer (BC) was the most frequently diagnosed cancer type among females worldwide (1–3), with more than 2 million new cases reported in 2020 (4) and over 300.000 additional cases recorded only in the USA in 2024 (5). Although breast cancer treatment is life-saving and essential, the disease and its treatment undeniably affect multiple aspects of the well-being of females with BC (see review: 69), including mental and physical health, as well as body image (6–8). All of these factors can collectively influence and worsen the quality of life of afflicted females (10–15).

Body image refers to the mental representation individuals form of their body, reflecting how they perceive their own physical appearance (16), and related concerns (17). It has both personal and interpersonal dimensions, involving (dis)satisfaction with one’s physical appearance, (diminished) femininity, and (compromised) body integrity, as well as beliefs about how others perceive oneself (18,19). BC diagnosis and its treatment can lead to a wide range of body-related side effects, including breast loss, changes in breast appearance, scarring, hair loss, weight and appetite fluctuations, treatment-induced menopause, potential infertility, sleep disturbances, musculoskeletal issues, and sexual health concerns such as reduced libido, vaginal dryness, and pain, all of which may negatively affect body image (17,20–23). Altered body image is a significant psychosocial concern for females with BC, with studies consistently reporting poorer body image compared to healthy controls (17,24–28). Ongoing treatment, in particular, has been linked to further declines in body image when comparing patients before and during treatment (29–32).

Given the pivotal role of treatment in shaping body image, it is essential to consider how different treatment modalities may differentially affect female’s perceptions of their bodies. Depending on tumor biology and cancer stage, treatment may include systemic therapy, radiotherapy, and surgery – either breast-conserving therapy (BCT) or mastectomy (MX) While BCT involves removal of the tumor and some surrounding tissue, MX entails complete removal of the breast. Studies comparing their effects on body image have yielded mixed results: some show more favorable outcomes with BCT (33–37), others report no significant differences (38,39), and one study even observed better body image in MX patients (26).

Recent literature has emphasized the need to advance our understanding of psychological adjustment in breast cancer by moving beyond symptom-specific or phase-limited perspectives toward integrative frameworks that capture mental health trajectories across the entire continuum of care. One such framework is the recently proposed Breast Cancer Psychological Integrative Phasic Model, which synthesizes existing theoretical and empirical work into a phase-sensitive structure encompassing screening, diagnosis, treatment, survivorship, and recurrence (43). Each phase is marked by distinct psychosocial challenges and associated symptom profiles, ranging from anxiety and trauma-related distress to depression and fear of recurrence, highlighting how mental health evolves in parallel with medical treatment. This phasic perspective provides a valuable lens for examining body image disturbances, which often arise or intensify during treatment and survivorship. Rather than treating body image problems in isolation, the model situates them within a broader constellation of psychological responses shaped by temporal and structural dynamics across the cancer trajectory.

Overall, BC and its treatment modalities appear to lead to mixed results regarding body image, and a quantitative evaluation of the effects of disease, different treatment modalities, and treatment durations is still lacking. To our knowledge, this is the first systematic review and meta-analysis to simultaneously examine body image changes in these three domains: 1) disease effect, 2) treatment effect, and 3) treatment modality. Therefore, a systematic review and meta-analysis of the existing data on body image in females with BC is required to offer a comprehensive overview of the field. This will help guide future studies, directing scientific inquiry to explore any gaps in understanding and improve knowledge for physicians, patients, and their relatives.

The aim of the study is to investigate:

1) Effect of disease (females with BC vs. healthy controls): How does the body image of females with BC differ from that of healthy individuals?
2) Effect of treatment (before vs. during): What changes occur in the body image of females with BC when comparing the before to the ongoing treatment phase?
3) Treatment modality (MX vs. BCT): How do body image perceptions, including both negative and positive aspects, vary between females with BC undergoing mastectomy (MX) and those receiving breast-conserving therapy (BCT)?

## 2. METHODS

A systematic review and meta-analysis on body image in females with BC was conducted according to the Preferred Reporting Items for Systematic Reviews and Meta-Analyses (PRISMA) 2020 statement (40). Prior to performing this study, a review protocol was registered in the International Prospective Register of Systematic Reviews (PROSPERO) database (Registration No. CRD42024503033; https://www.crd.york.ac.uk/prospero/display_record.php?ID=CRD42024503033).

### 2.1 Search strategy

Published studies up to 2025 were searched in PubMed, APA PsycINFO, CINAHL, and Web of Science Core Collection by Uppsala University Library. The following search terms were used: (“breast neoplasm*” OR “breast cancer*” OR “breast tumor*” OR “breast carcinoma*” OR “mammary cancer*” OR “mammary carcinoma*” OR “mammary tumor*” OR “mammary neoplasm*”) AND (“female*” OR “girl*” OR “woman*” OR “women*”) AND (“body image” OR “body acceptance” OR “body schema” OR “body dissatisfaction” OR “body esteem” OR “body image disturbance*” OR “body checking” OR “dysphoria” OR “physical self” OR “self-image” OR “body shame” OR “body perception” OR “body satisfaction” OR “body appreciation”)

### 2.2 Eligibility criteria

Articles were considered eligible if they met the following inclusion criteria: (1) original research; (2) measuring body image as a primary outcome variable (a full list of questionnaires is provided in Supplementary Table 1); (3) fully accessible text; (4) studies on female breast cancer patients vs. healthy controls, baseline vs. ongoing breast cancer treatment process, MX vs. BCT; (5) studies only cover current female breast cancer patients with no metastasis and no breast reconstruction; (6) observational studies. Articles were excluded when they met one of the following criteria: (1) non-peer-reviewed manuscripts, non-research articles, non-English articles, meeting abstracts; (2) studies on physical, psychological, or psychosocial intervention/counseling; (3) studies on couple-based issues; (4) studies on unmet needs; (5) lack of a control group.

This study included only females with non-metastatic BC to increase validity and consistency, as metastatic BC occurs when cancer has spread beyond the breast, indicating an advanced stage, and is often associated with severe symptoms such as fatigue and pain, which can exacerbate treatment side effects (41,42).

### 2.3 Study selection and Data extraction

Two reviewers (SYS and RW) independently extracted relevant data from the included studies. Any discrepancies were identified and resolved through consensus in consultation with co-authors MH and BD. The following information was extracted for all articles: (1) Publication information (title/author/year/journal); (2) Study characteristics (aim/study design/data collection/mean and standard deviation); (3) Participants (sample size/age/treatment/symptom); (4) Outcomes (instrument-related data, mean results).

In the meta-analysis, mean and standard deviation values were directly extracted from the original tables in the studies. Transforming scale orientations (e.g., for positive vs. negative body image) was avoided to preserve data integrity, as such transformations could compromise the original meaning and reliability of the data reported by the primary sources. Various body image-related questionnaires were included in the meta-analysis (see Supplementary Table 1). Positive body image was defined by higher scores on relevant questionnaires, whereas negative body image was indicated by higher scores on specific questionnaires. As such, two separate meta-analyses were conducted for the comparison of MX vs BCT groups.

If any articles included two groups/subgroups, data were extracted separately for each group/subgroup. When the study measured body image repeatedly (more than two times) in a longitudinal fashion, we included only two distinct measurement time points, each with a larger number of participants, to minimize the risk of result duplication and ensure the robustness of our analysis. When multiple studies included the same sample, only the study with the largest sample was included to avoid duplication of participants. Articles that described body image but lacked quantitative data – despite attempts to obtain this information from the authors via email – were included in the descriptive systematic review rather than the quantitative meta-analysis.

### 2.4 Quality assessment

#### 2.4.1 Assessment of risk of bias

The risk of bias for each included paper was evaluated independently by two reviewers using the Newcastle-Ottawa Scale (NOS) (43) and the Risk Of Bias in Non-randomized Studies of Interventions Scale (ROBINS-I) (44) based on the study characteristics. The NOS is a straightforward quality assessment tool specifically for non-randomized cohort and case-control studies, focusing on three elements: selection of study groups, comparability between groups, and allocation of the exposure or outcome of interest. Assessment quality is categorized as low (0–4), moderate (5–7), or high (8–10). For some studies, it was not possible to apply the NOS, in these cases the ROBINS-I tool was applied. The ROBINS-I is a comprehensive tool suitable for various study designs, offering a nuanced quality assessment through detailed questions on potential biases, such as those in reported outcomes and intervention classification. Specifically designed for non-randomized intervention studies, ROBINS-I evaluates seven bias elements: confounding, participant selection, intervention classification, deviations from intended interventions, missing data, outcome measurement, and selection of reported outcomes. Quality is rated in four categories: low, moderate, serious, and critical.

#### 2.4.2 Publication bias

The publication bias was assessed by visual inspection of funnel plots and Egger’s regression test for asymmetry (45) using the JAMOVI (2.3.38) software (46). Significant publication biases were identified by p-values lower than 0.05.

### 2.5 Statistical analysis

We analyzed the differences in body image between (1) females with BC vs. healthy controls, (2) females with BC before vs. during treatment, and (3) females with BC who had undergone MX vs. BCT. The standardized mean differences and their 95% confidence intervals (CI) were calculated using a Random Effects (RE) model with between-study variance estimated via Restricted Maximum Likelihood (REML) (47), due to substantial heterogeneity in clinical samples and methodological approaches between studies. Mean, standard deviation and sample size were provided from the included studies or corresponding authors. All statistical analyses were performed using JAMOVI (version 2.3.38) (46), and the alpha level was set at p<0.05. Forest plots present the overall effect size estimates along with their corresponding 95% confidence intervals. The number of included studies in each analysis was reported by “k” and effect size was reported by “μ”. Heterogeneity among the studies in each analysis was evaluated using Cochran’s Q and Higgins’ I^2^ statistics (43). A threshold of 25%, 50%, and 75% was used to differentiate between low, medium, and high heterogeneity.

## 3. RESULTS

### 3.1 Literature retrieval results and study characteristics

The database search initially yielded a total of 6738 studies. Based on the titles and abstracts, the first screening excluded 3904 studies and retained 363 studies for further evaluation. After the full-text screening, 322 studies were excluded. See Fig 1 for a flow chart of article selection following the PRISMA 2020 guidelines.

**Fig 1.**
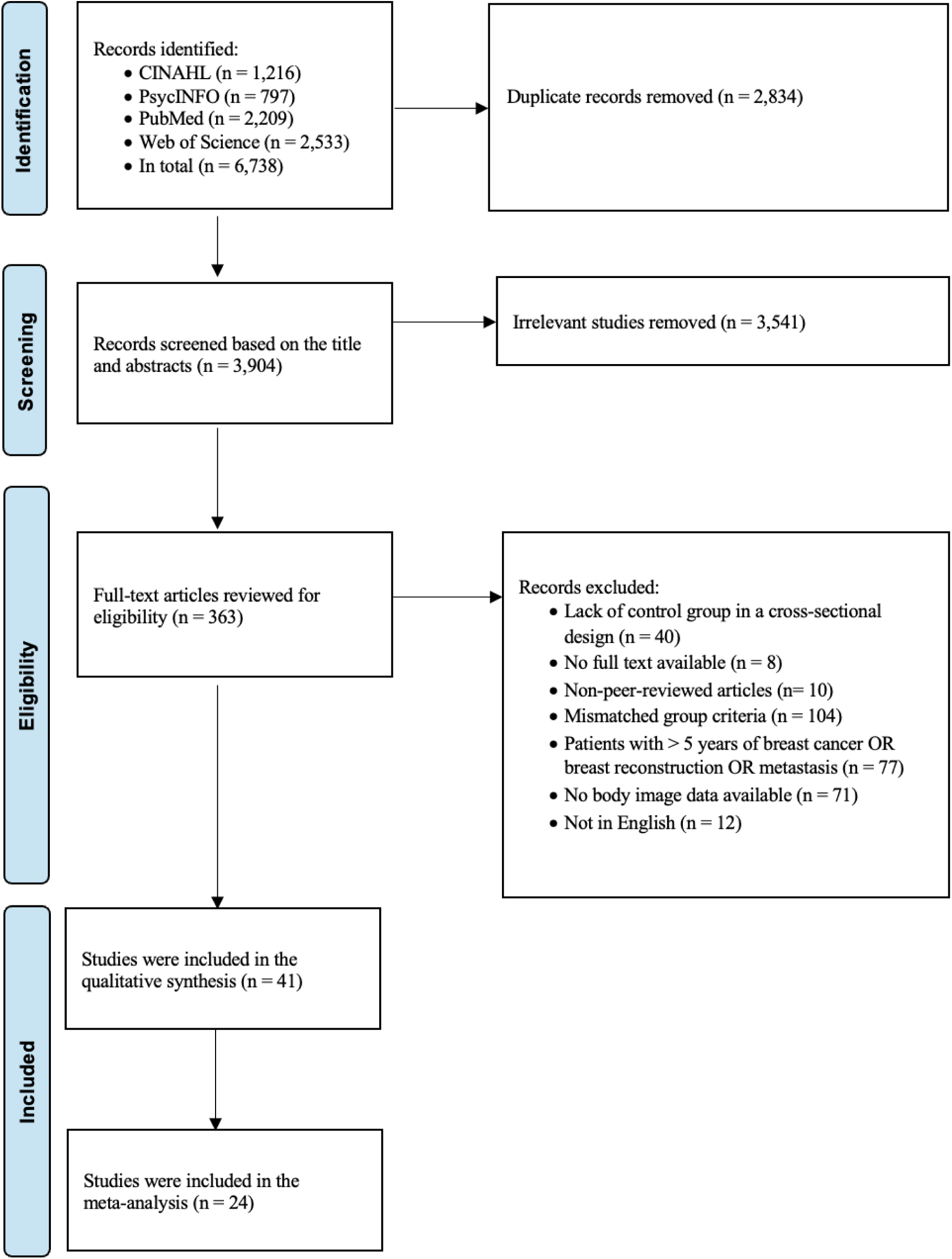
PRISMA flow diagram of the study search and selection process.

Ultimately, 41 studies were included in the final analysis of this study. The included studies were published between 1988 and 2025. Based on the availability of body image scores, 24 body image-related studies contained adequate data for quantitative synthesis in the meta-analysis. Five reported differences between females with BC and controls, ten compared before vs. during treatment, six compared MX vs. BCT in the negative body image direction, and eight estimated MX vs. BCT in the positive body image direction. Some studies contributed data to multiple meta-analyses due to their design and reporting structure. Specifically, certain studies included both cross-sectional comparisons (e.g., females with breast cancer vs. healthy controls) and longitudinal assessments (e.g., pre-treatment vs. during treatment) or provided subgroup analyses based on surgical type (e.g., MX vs. BCT). For instance, a single study might compare females with BC to healthy controls while also reporting within-group changes over time among the breast cancer cohort, thereby contributing to a second meta-analysis. There were 4 studies that were included in more than one meta-analysis. Those studies marked in Table 1-3 to indicate which data were used for which meta-analysis. These studies included 6872 females with BC and 398 healthy controls. The general characteristics of the 41 included studies are presented in Tables 1-3 with the studies that could not be included in the meta-analysis highlighted with asterisks.

**Table 1:** Characteristics of included studies investigating body image (BI) in female with breast cancer (BCP) and healthy controls (HC).

| Study | CO | BCP |  |  |  |  | HC |  |  | Matched<br>by age | BI Qu. | Quality of<br>study |
| --- | --- | --- | --- | --- | --- | --- | --- | --- | --- | --- | --- | --- |
|  |  | N | Age<br>(mean/<br>range) | BI score,<br>mean<br>(SD) | Tumor size/<br>Cancer stage | Treatment | N | Age<br>(mean/<br>range) | BI score,<br>mean<br>(SD) |  |  |  |
| Aerts et al., 2014 | BE | 56 | 57/<br>34-75 | 10.54<br>(8.17) | na/<br>Early BC stage | Surgery,<br>Chemotherapy,<br>Radiation therapy,<br>Endocrine therapy | 81 | 56/<br>26-80 | 4.49<br>(5.04) | Yes | BIS | NOS: 7 |
| Kraus, 1999 | USA | 31 | 52/<br>29-82 | 122.94<br>(3.33) | na/<br>Early BC stage | Surgery | 30 | 53/<br>29-82 | 116.47<br>(3.30) | na | BIS* | NOS: 4 |
| Montañés Muro et al.,<br>2023 | SPA | 27 | 54/<br>38-70 | 0.78<br>(0.44) | na/<br>na | Surgery | 27 | 50/<br>37-62 | 0.50<br>(0.27) | na | BIS | NOS: 7 |
| Savani et al., 2023 | IND | 60 | 46/<br>na | 10.48<br>(9.23) | na/<br>na | na | 60 | 46/<br>na | 1.60<br>(2.49) | Yes | BIS | NOS: 7 |
| Jahromi et al., 2022 | IRI | 111 | 48/<br>30-55 | 51.56<br>(15.09) | Cancer grade<br>2 | Chemotherapy,<br>Radiotherapy, Surgery | 111 | 47/<br>30-55 | 45.80<br>(16.82) | Yes | FOBIQ | NOS: 8 |
| *Bagheri & Mazaheri,<br>2015 | IRI | 50 | 32/<br>na | 109.94<br>(19.01) | na/<br>na | Radiotherapy | 50 | 33/<br>X | 120.20<br>(19.82) | na | MBSRQ | NOS: 7 |
| *Jablonski et al., 2018 | PL | 39 | 55/<br>34-68 | na | na | Surgery, BCT | 39 | 52/<br>34-68 | na | na | BS-Q | NOS:7 |
| *Izdorczyk et al., 2018 | PL | 120 | 55/<br>na. | na | na | Mastectomy | 99 | 54/<br>n.a. | na | Yes | BAQ | NOS:4 |
| *Anagnostopolous & Myrghianni, 2009 | GR | 70 | na | na | na | Surgery,<br>Chemotherapy,<br>Endocrine therapy,<br>Radiotherapy | 70 | n.a. | na | na | BIS | NOS:7 |

*Note.* SD: Standard deviation; BIS: Body Image Scale (96); BIS\*: Body Image Scale- Modified version (97); MBSRQ: Multidimensional Body-Self Relations Questionnaire (98); FOBIQ; Fear of Body Image Questionnaire (99); BS-Q: Body self-questionnaire (Mirucka, 2017); BAQ: Body Attitude Questionnaire (100) ; na.: Missing data; NOS: Newcastle-Ottawa scale. BCT: Breast conservative therapy; BI QU: Body Image related questionnaires; \*: Studies in only Qualitative analysis; CO: Country; GR: Greece; PL:Poland; IRI: Iran; SPA: Spain; BE: Belgium; IND: India.

The geographic distribution of the studies included in the systematic review and meta-analysis is depicted in a pie chart (Fig. 2), highlighting potential cultural influences on body image among females with BC.

**Fig 2.**
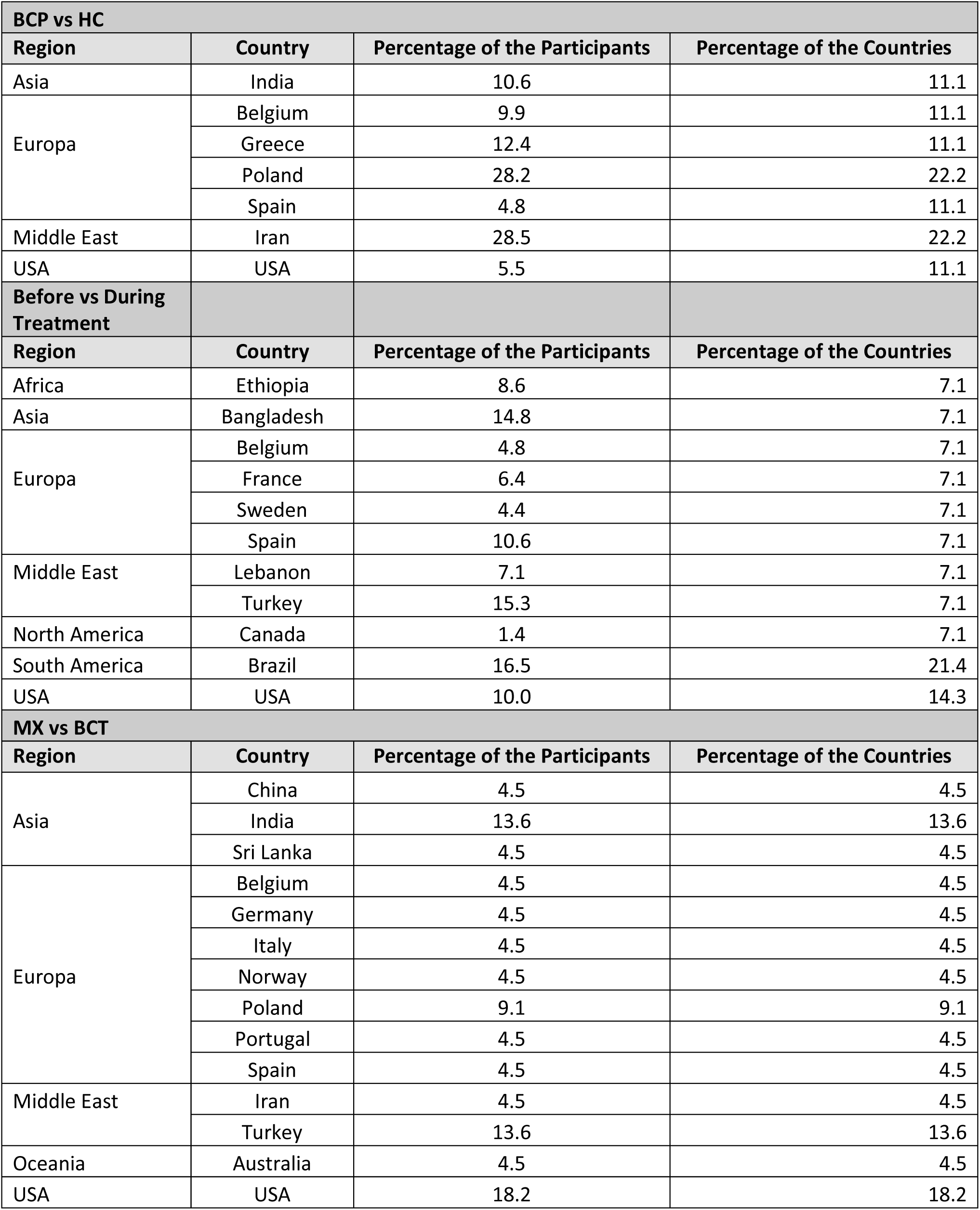
The table illustrates the percentage of countries and samples of each country included in the study

### 3.2 Meta-analyses & Qualitative analyses

#### 3.2.1 Comparison of body image in Females with BC vs. Healthy Controls

Nine articles were identified targeting this comparison and were published between 1999 to 2023. Among them, four studies lacked quantitative body image data and thus were included in the systematic review (see Table 1 for their characteristics).

Five studies were included in this meta-analysis and reported data from 469 females with BC and 479 healthy controls (with three studies using age-matched controls). They were conducted in Belgium, the USA, Spain, Iran, and India (17,26–28,48).

Compared to healthy controls, females with BC had a significantly worse body image score (k = 5, μ =-1.02 95 %CI [-1.53,-0.51], p < 0.001, see Fig 3). This finding presented a large effect size in the negative direction (ES =-1.02). The heterogeneity between studies was high (I^2^ = 87.23%, p < 0.001).

**Fig 3:**
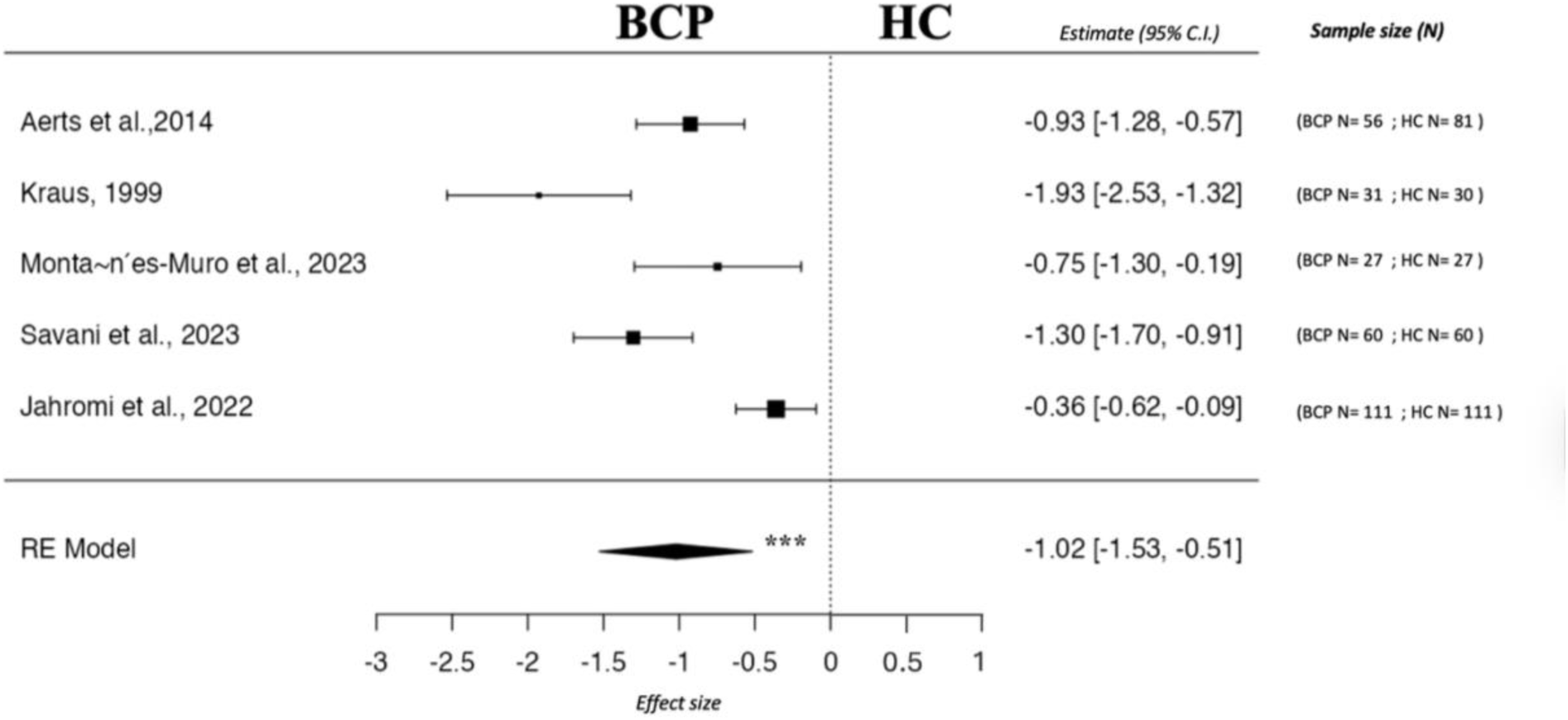
Forest plot of body image scores comparing values from females with breast cancer (BCP) and healthy controls (HC). Note. *** marks significant group difference with p < 0.001. Abbreviations: RE= Random Effects

##### 3.2.1.2 Qualitative analysis (females with BC vs. healthy controls)

Studies report significantly higher levels of body dissatisfaction in females with BC than in healthy controls (24,25,49). Additionally, lower body acceptance was shown in females with BC compared to healthy controls (50).

#### 3.2.2 Comparison of body image before vs during treatment

Fourteen articles were identified targeting this comparison and were published between 1998 to 2023. Among them, four studies lacked quantitative body image data and thus were included only in the systematic review (see Table 2 for their characteristics).

**Table 2:** Characteristics of included studies investigating body image (BI) in baseline and during treatment females with breast cancer (BC).

| Study | CO | Before treatment |  | During treatment |  | Age (mean/<br>range) | Tumor size/<br>cancer stage | Treatment | BI QU | Quality of study |
| --- | --- | --- | --- | --- | --- | --- | --- | --- | --- | --- |
|  |  | N | BI score, mean (SD) | N | BI score, mean (SD) |  |  |  |  |  |
| Aerts et al., 2014 | BE | 81 | 1.53 (3.48) | 66 | 4.05 (5.52) | 57/<br>34-75 | na./<br>Early stage BC | Surgery,<br>Chemotherapy,<br>Radiotherapy,<br>Endocrine therapy | EORTC QOL-BR23 | NOS: 6 |
| Bueno et al., 2018 | BR | 166 | 84.50 (23.40) | 166 | 80.60 (26.80) | 59/<br>na | na./<br>na | Surgery | EORTC QOL-BR23 | NOS: 4 |
| Browall et al., 2008 | SE | 75 | 90.00 (18.00) | 65 | 66.00 (30.00) | 64/<br>55-80 | na./<br>Stage I:<br>23<br>Other stages:<br>52 | Chemotherapy,<br>Radiotherapy,<br>Endocrine therapy, Surgery | EORTC QOL-BR23 | NOS: 6 |
| Rahman et al., 2015 | BD | 250 | 86.00 (7.83) | 250 | 70.46 (6.36) | 44/<br>21-67 | T1: 15<br>T2: 102<br>T3: 53<br>T4: 80/<br>na. | Mastectomy, Neoadjuvant chemotherapy | EORTC QOL-BR23 | NOS: 8 |
| Viller et al., 2017 | SPA | 180 | 94.20<br>(13.20) | 180 | 85.10<br>(23.00) | 58/<br>na | na./<br>0 : 22<br>I : 149<br>II : 128<br>III : 35<br>IV: 2 | Chemotherapy,<br>Radiotherapy,<br>Hormone therapy,<br>Biological therapy anti-Her2 | EORTC QOL-BR23 | NOS: 8 |
| Binotto et al., 2020 | BR | 33 | 90.66<br>(14.99) | 33 | 66.41<br>(31.49) | 51/<br>na. | na./<br>Stage 1, 2 and<br>3 | Surgery,<br>Adjuvant or Neoadjuvant<br>Taxane-based<br>chemotherapy | EORTC QOL-BR23 | NOS: 6 |
| Castro e Silva et al.,<br>2021 | BR | 80 | 25.73<br>(18.57) | 80 | 34.06<br>(6.10) | 52/<br>X | na./<br>Any at stage | Surgery,<br>Chemotherapy | EORTC QOL-BR23 | NOS: 5 |
| Bahar et al., 2023 | TUR | 259 | 87.00<br>(18.00) | 197 | 94.00<br>(13) | 51/<br>24-84 | na./<br>I: 95<br>II: 156<br>III: 206 | Radiotherapy,<br>Surgery, Chemotherapy,<br>Hormone therapy | EORTC QOL-BR23 | NOS: 5 |
| El Haidari et al., 2023 | LB | 120 | 89.20<br>(17.70) | 116 | 81.00<br>(21.60) | 54/<br>na. | T1: 42<br>T2: 71<br>missing: 7/<br>I: 52<br>II: 68 | Surgery | EORTC QOL-BR23 | NOS: 5 |
| *Parker et al., 2007 | USA | 104 | 23.80<br>(5.50) | 104 | 22.60<br>(6.10) | 53/<br>na | na./<br>stage I and II | Chemotherapy,<br>Radiotherapy,<br>Surgery | EORTC QOL-BR23 | NOS: 8 |
| *Gadisa et al., 2021 | ET | 146 | na | na | na | 42/<br>na | I: 6<br>II: 48<br>III: 64<br>IV: 28 | Chemotherapy | EORTC QOL-BR23 | NOS: 7 |
| *Laroche et al., 2017 | FR | 109 | 66.00<br>(33) | 109 | na | 61/<br>na. | Early stage BC | Surgery,<br>Chemotherapy, Aromatase<br>inhibitor | EORTC QOL-BR23 | NOS: 4 |
| *Carver et al., 1998 | USA | 66 | na | 61 | na | 52/<br>28-76 | I: 49<br>II: 17 | Surgery,<br>Chemotherapy, Hormone<br>therapy, Radiotherapy | MBA | NOS: 3 |
| *Metcalfe et al., 2012 | CA | 24 | na | 24 | na | 53/<br>30-83 | 0: 19<br>I: 26<br>II: 41<br>III: 14 | Surgery,<br>Radiotherapy | BIBC | NOS: 5 |

*Note.* SD: Standard deviation; BIS: Body Image Scale (96) ; BIS\*: Body Image Scale- Modified version (97); EORT QoL-BR 23: European Organization for Research and Treatment of Cancer quality of life in patients with breast cancer (101); BIBC: Body image after breast cancer (Baxter, 1997); MBA: Measure of Body Apperception (30); NOS: Newcastle-Ottawa scale; na.: Missing data, BI QU: Body Image related questionnaires; \*: Studies in only Qualitative analysis; CO: Country ; CA: Canada; FR: France; ET: Ethiopia; LB: Lebanon; TUR: Turkey; BR: Brazil; SPA: Spain; BE: Belgium; BD: Bangladesh; SE: Sweden.

**Table 3:** Characteristics of included studies investigating body image (BI) in females with breast cancer (BCP) treated with breast-conserving therapy (BCT) and mastectomy (MX).

| Study | CO | BCP with BCT |  |  |  | BCP with MX |  |  |  | Age<br>(mean<br>/<br>range) | BI QU | Quality<br>of study |
| --- | --- | --- | --- | --- | --- | --- | --- | --- | --- | --- | --- | --- |
|  |  | N | BI score<br>mean<br>(SD) | Tumor size/<br>Cancer stage | Treatment | N | BI score<br>mean<br>(SD) | Tumor size/<br>Cancer stage | Treatment |  |  |  |
| <sup>a</sup>Jendrian et al., 2017 | GER | 46 | 6.84<br>(6.30) | Tis/T1: 36<br>T2/T3/T4: 5 | Surgery,<br>Chemotherapy,<br>Endocrine therapy | 61 | 11.51<br>(7.67) | Tis/T1: 44<br>T2/T3/T4: 12 | Surgery,<br>Chemotherapy,<br>Endocrine therapy | 68/<br>36.4-83.8 | BIS | NOS: 6 |
| <sup>b</sup>Jayasinghe et al., 2021 | SL | 19 | 58.35<br>(55.39) | T1: 15<br>T2: 27<br>T3 :5<br>T4 :2 | Surgery,<br>Adjuvant<br>Chemotherapy,<br>Hormone therapy,<br>Adjuvant<br>Radiotherapy,<br>Axillary surgery | 35 | 62.51<br>(27.68) | T1: 15<br>T2: 27<br>T3: 5<br>T4: 2 | Surgery,<br>Chemotherapy,<br>Hormone therapy,<br>Radiotherapy,<br>Axillary surgery | 58/<br>36-81 | EORT QoL-BR 23 | ROBINS-I:<br>MODERATE |
| <sup>b</sup>Munshi et al., 2010 | IND | 142 | 77.70<br>(21.80) | Early BC: 119<br>Advanced BC: 23 | Adjuvant<br>radiotherapy,<br>Surgery | 113 | 79.80<br>(35.20) | Early BC: 51<br>Advanced BC: 62 | Adjuvant<br>radiotherapy,<br>Surgery | na | EORT QoL-BR 23 | ROBINS-I:<br>LOW |
| <sup>b</sup>King et al., 2000 | AU | 261 | 71.00<br>(15.00) | T1 and T2 (no<br>exact number)/<br>Early stage of BC | Surgery,<br>Radiotherapy,<br>Chemotherapy | 43 | 48<br>(28) | T1 and T2 (na)/<br>Early stage of<br>BC | Surgery,<br>Radiotherapy,<br>Chemotherapy | BCT:54<br>/25-81<br>MX:53 | ESBC | NOS: 5 |
| <sup>b</sup>Konieczny and Fal,<br>2023 | PL | 125 | 58.97<br>(32.22) | na. | Surgery,<br>Radiotherapy,<br>Chemotherapy,<br>Hormonal therapy,<br>Systemic therapy | 64 | 58.73<br>(31.16) | na. | Surgery,<br>Radiotherapy,<br>Chemotherapy,<br>Hormonal therapy,<br>Systemic therapy | 55/<br>na. | EORT QoL-BR 23 | ROBINS-I:<br>LOW |
| <sup>b</sup>Dubashi et al.,<br>2010 | IND | 18 | 76.88<br>(33.03) | T1: 2<br>T2: 31<br>T3: 18 | Surgery,<br>Chemotherapy,<br>Hormonal therapy,<br>Bilateral salpingo-<br>oophorectomy | 33 | 82.38<br>(26.20) | T1: 2<br>T2: 31<br>T3: 18 | Surgery,<br>Chemotherapy,<br>Hormonal therapy,<br>Bilateral salpingo-<br>oophorectomy | 30/<br>na. | EORT QoL-BR 23 | ROBINS-I:<br>LOW |
| <sup>b</sup>Mock, 1988 | USA | 90 | 97.20<br>(12.31) | Early BC stage | Surgery,<br>Adjuvant<br>chemotherapy | 62 | 98.20<br>(12.47) | Early BC stage | Surgery,<br>Adjuvant<br>chemotherapy | 52/<br>29-79 | BIS | NOS: 4 |
| <sup>b</sup>Jankowska-Polańska et al.,<br>2020 | PL | 50 | 74.50<br>(24.99) | na./<br>na | Surgery,<br>Radiotherapy,<br>Chemotherapy,<br>Hormonal therapy | 50 | 57.39<br>(24.25) | na./<br>na | Surgery,<br>Radiotherapy,<br>Chemotherapy,<br>Hormonal therapy | 53/<br>na | EORT QoL-BR 23 | ROBINS-I:<br>LOW |
| <sup>a</sup>Ardakani et al., 2020 | IRI | 109 | 12.87<br>(6.41) | T1: 10<br>T2: 11<br>T3: 8<br>T4: 11<br>na: 140<br>na | Surgery,<br>Radiotherapy,<br>Chemotherapy,<br>Hormonal therapy | 71 | 15.18<br>(6.51) | T1: 10<br>T2: 11<br>T3: 8<br>T4: 11<br>na: 140<br>na | Surgery,<br>Radiotherapy,<br>Chemotherapy,<br>Hormonal therapy | 46/<br>25-70 | BIS | ROBINS-I:<br>LOW |
| <sup>a</sup>Fonseca et al., 2014 | PRT | 26 | 3.69 | na./<br>na | Surgery,<br>Neoadjuvant<br>chemotherapy | 29 | 8.24<br>(7.22) | na./<br>na | Surgery,<br>Neoadjuvant<br>chemotherapy | 54/<br>32-79 | BIS | ROBINS-I:<br>MODERATE |
| <sup>a</sup>Aerts et al., 2014 | BE | 81 | 1.53<br>(3.48) | na./<br>na. | Surgery,<br>Radiotherapy,<br>Chemotherapy,<br>Hormonal therapy | 68 | 6.99<br>(7.58) | na./<br>na. | Surgery,<br>Radiotherapy,<br>Chemotherapy,<br>Hormonal therapy | BCT:<br>57/<br>34-75<br>MX:<br>54/<br>26-74 | BIS | NOS: 7 |
| Montañés -Muro et al., 2023 | SPA | 13 | 0.64<br>(0.38) | na./<br>na | Surgery | 14 | 0.91<br>(0.47) | na./<br>na. | Surgery | 53/<br>38-70 | BIS | NOS: 7 |
| <sup>b</sup>Parker et al., 2007 | USA | 104 | 23.6<br>(6.90) | na./<br>Stage I and II BC | Chemotherapy,<br>Radiotherapy,<br>Surgery | 45 | 22.50<br>(5.10) | na./<br>Stage I and II<br>BC | Chemotherapy,<br>Radiotherapy,<br>Surgery | BCT :<br>53<br>MX:<br>52/<br>na | MBSRQ | NOS: 8 |
| *Shekhar et al., 2023 | IND | 402 | 2.78<br>(3.67) | T1: 215<br>T2: 187/<br>0: 10<br>1: 72<br>2: 253<br>3: 34 | Hormone therapy,<br>Radiotherapy,<br>Surgery | 304 | 29.38<br>(11.55) | T2: 148<br>T3: 156/<br>0: 24<br>3: 280 | Hormone therapy,<br>Radiotherapy,<br>Surgery | < 35 :<br>28<br>35-55:<br>362<br>55-70:<br>266<br>>70:<br>50 | OBC | ROBINS-I:<br>LOW |
| *Spatuzzi et al.,2016 | ITA | 72 | na | BC stage 1-2 | Surgery,<br>Chemotherapy,<br>Radiotherapy,<br>Hormone therapy | 44 | na | BC stage 1 & 2 | Surgery,<br>Chemotherapy,<br>Radiotherapy,<br>Hormone therapy | BCT<br>mean:<br>57<br>MX<br>mean:<br>61.3 | EORT QoL-BR 23 | ROBINS-I:<br>LOW |
| *Arıcan Alicikus et al., 2009 | TUR | 61 | na | na | Surgery,<br>Chemotherapy,<br>Radiotherapy,<br>Hormone therapy | 51 | na | na | Surgery,<br>Chemotherapy,<br>Radiotherapy,<br>Hormone therapy | 50/<br>31-65 | MBSRQ | ROBINS-I:<br>LOW |
| *Yilmazer et al., 1994 | TUR | 40 | na | BC stage 1& 2 | Surgery,<br>Chemotherapy,<br>Radiotherapy | 40 | na | BC stage 1 and 2 | Surgery,<br>Chemotherapy,<br>Radiotherapy | BCT:<br>40/<br>27-50<br>MX:<br>43/<br>30-50 | A draw a person test | ROBINS-I:<br>MODERATE |
| *Fung et al., 2001 | CHN | 17 | na | Tis:1<br>T1:10<br>T2:6 | Surgery | 15 | na | Tis:1<br>T1: 4<br>T2:!0 | Surgery | BCT:<br>47<br>/na<br>MX:<br>50<br>/na | Adopted initially<br>the body image<br>index | ROBINS-I:<br>LOW |
| *Schou et al., 2005 | NOR | 85 | na | Tis:2<br>T1:68<br>T2:30 | Systematic<br>therapy,<br>Surgery,<br>Radiotherapy | 66 | na | Tis:2<br>T1:68<br>T2:30 | Systematic<br>therapy, Surgery,<br>Radiotherapy | 56/<br>21-78 | CAWAC | ROBINS-I:<br>LOW |
| *Yurek et al., 2000 | USA | 78 | na | T2: 165<br>T3: 25 | Surgery,<br>Adjuvant therapy | 79 | na | T2: 165<br>T3: 25 | Surgery,<br>Adjuvant therapy | 51/<br>30-84 | BSS | ROBINS-I:<br>LOW |
| *Arikan et al., 2020 | TUR | 52 | na | T1:9<br>T2: 71<br>T3: 19<br>T4: 18 | Surgery,<br>Chemotherapy,<br>Target therapy,<br>Hormone therapy | 58 | na | T1:9<br>T2: 71<br>T3: 19<br>T4: 18 | Surgery,<br>Chemotherapy,<br>Target therapy,<br>Hormone therapy | 56/<br>27-91 | BIS | ROBINS-I:<br>LOW |
| *Afshar-Bakshloo et al.,2023 | GER | 212 | Na | T0: 161<br>T1: 48 | Surgery,<br>Chemotherapy,<br>Target therapy,<br>Radiotherapy | 27 | Na | T0: 27 | Surgery,<br>Chemotherapy,<br>Target therapy,<br>Radiotherapy |  | EORT QoL-BR 23 | ROBINS-I:<br>MODERATE |
| *Kraus, 1999 | USA | 16 | na | Early BC | na | 15 | na | Early BC | Surgery | 52/<br>29-82 | BIS* | NOS: 4 |

*Note.* BIS: Body Image Scale (96) ; BIS\*: Body Image Scale- Modified version (97); OBC: Objectified body consciousness (102); MBSRQ: Multidimensional Body-Self relations questionnaire (98) ; EORT QoL-BR 23: European Organization for Research and Treatment of Cancer quality of life in patients with breast cancer (101); CAWAC: Caring about women and cancer (103);

Ten studies were included in this meta-analysis reporting data from 1262 females with BC before treatment and 1245 females with BC during treatment. The studies were conducted in Belgium, Turkey, Brazil (three studies), Sweden, Lebanon, Spain, the USA and Bangladesh (48,51–59).

Comparing body image data before vs during treatment, the meta-analysis revealed that patients had significantly higher body image before the start of treatment than during treatment (k = 13 μ = 0.449, 95 %CI [-0.83,-0.07], p = 0.019, see Fig 4). This finding presented a small to moderate negative effect size (ES =-0.45). The heterogeneity between studies was high (I^2^ = 95.04%, p < 0.001).

**Fig 4:**
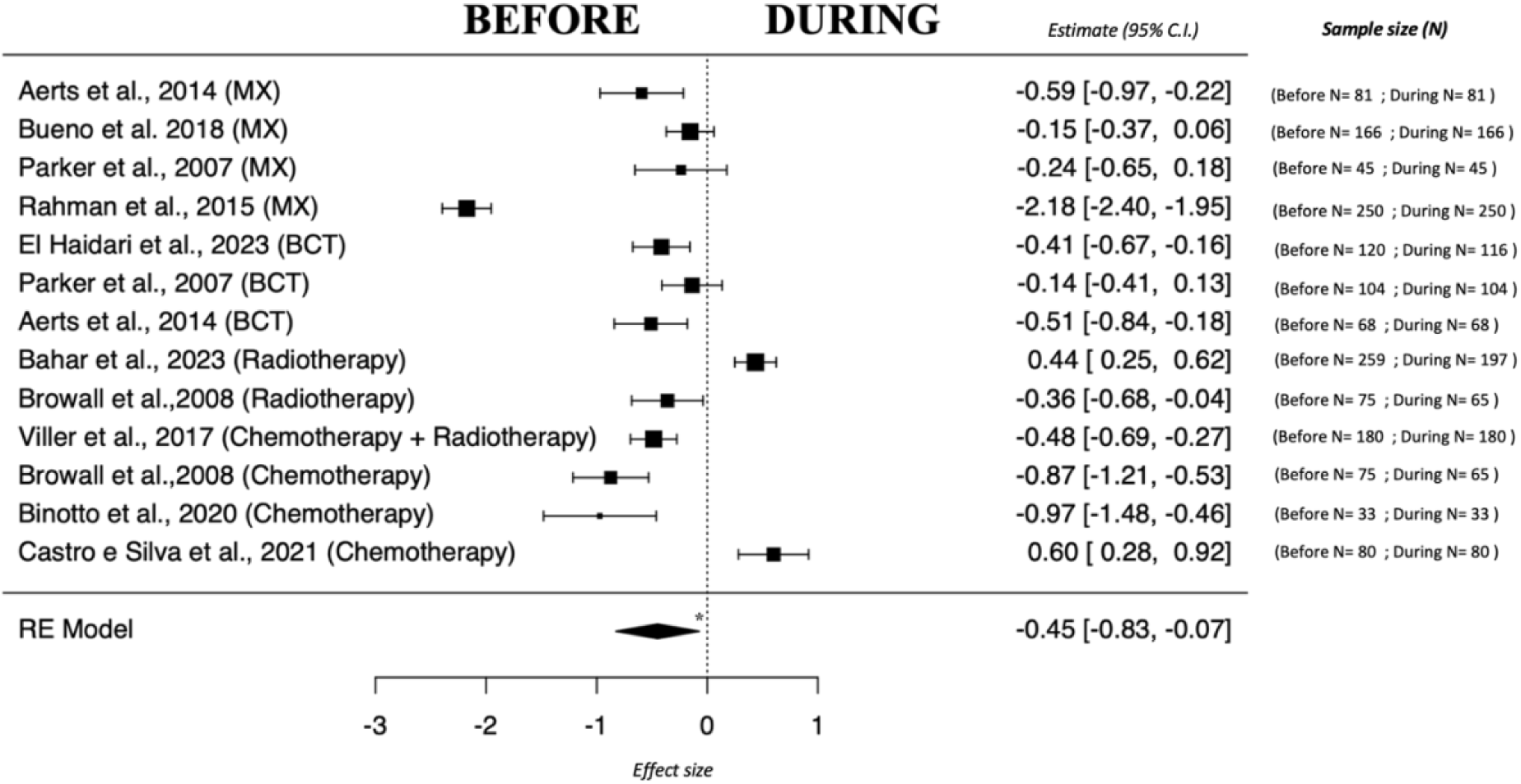
Forest plot of body image for the comparison between before vs. during treatment. Note. Treatment modalities are included in this forest plot. MX = mastectomy, BCT = breast-conserving therapy, * marks significant group difference with p < 0.05 Abbreviations: RE= Random Effects

##### 3.2.2.1 Qualitative analysis (Before vs. during treatment)

Three additional studies show that females with BC have a negative body image during treatment than before treatment (30–32). However, one study (60) reported no significant change in body image before vs. during breast cancer treatment.

#### 3.2.3 Comparison of MX vs BCT- negative and positive direction

Twenty-two articles were identified targeting this comparison and were published between 1998 to 2023. Among them, eight studies lacked quantitative body image data and thus were included only in the systematic review (see Table 3 for their characteristics).

##### 3.2.3.1 Meta-analysis of negative direction

Six studies were included in this meta-analysis that measured the negative direction of body image and relied on data from 677 BCT and 547 MX. They were conducted in Belgium, Iran, Portugal, Germany, Spain, and India (27,48,61–64).

Compared to the BCT group, the MX group had significantly higher negative body image (k = 6, μ =-1.105, 95 %CI [-2.01,-0.20], p = 0.016, see Fig 5a). This finding presented a large effect size in the negative direction (ES =-1.11). Again, the heterogeneity between studies was high (I^2^ = 97.19%, p < 0.001).

**Fig 5a&b:**
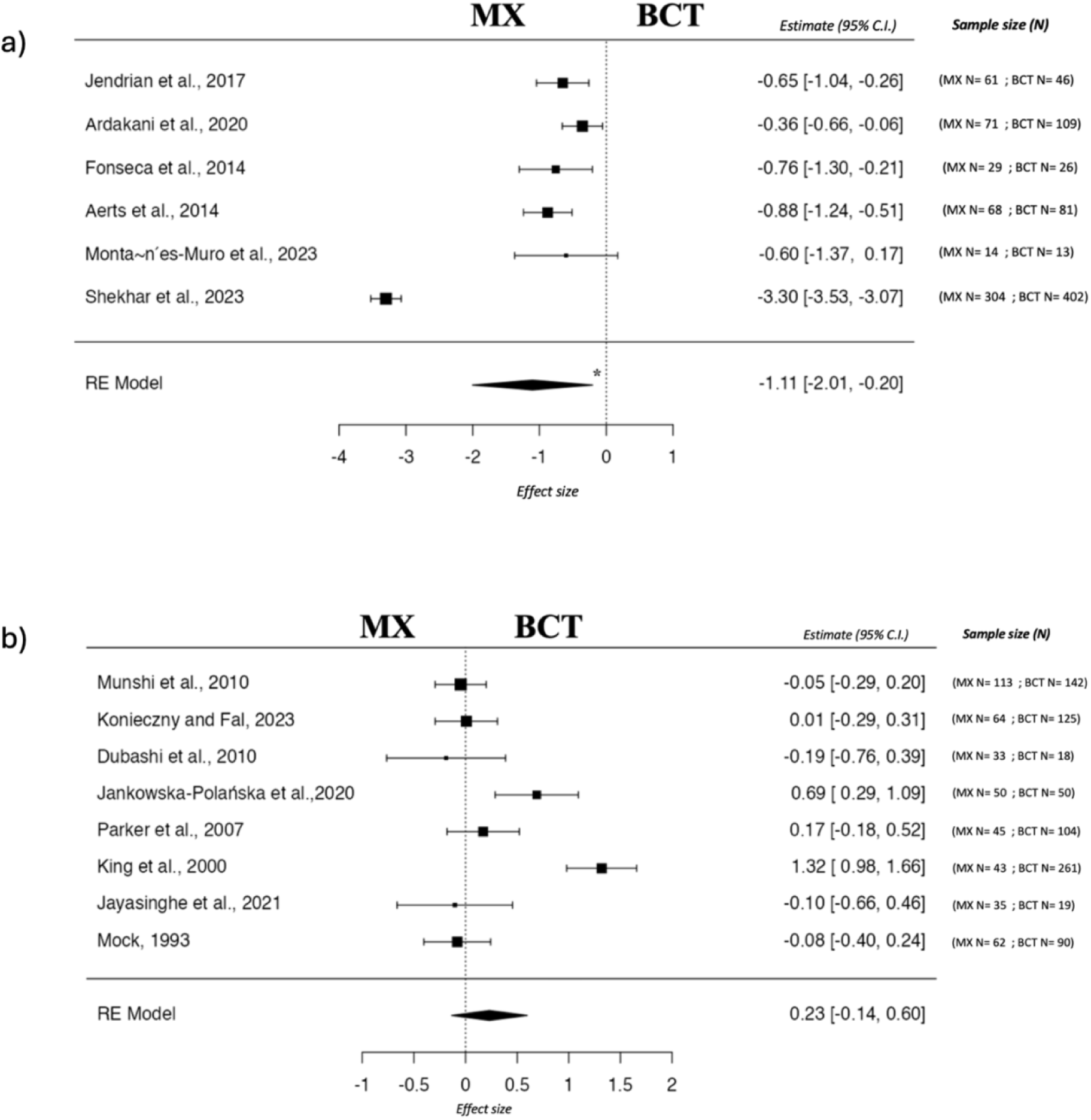
a) Forest plot of negative body image for the comparison between mastectomy (MX) and breast conservative therapy (BCT) Note. Negative body image refers to a person’s dissatisfaction with their physical appearance or the way they perceive their body, * marks significant group difference with p < 0.05. b) Forest plot of positive body image for the comparison between mastectomy (MX) and breast conservative therapy (BCT). Note. Positive body image means having a satisfactory and accepting attitude towards one’s body and appearance. Abbreviations: RE= Random Effects

##### 3.2.3.2 Meta-analysis of positive direction

Eight studies were included in this meta-analysis that measured the positive direction of body image and reported data from 809 BCT and 445 MX. Studies were conducted in India (two studies), Poland (two studies), Sri Lanka, Australia, and the USA (two studies) (57,65–71).

The meta-analysis revealed no significant difference between the BCT group and MX group, i.e., there was no group difference in terms of having a satisfactory and accepting attitude towards body image and appearance (k = 8, μ = 0.231, 95 %CI [-0.14, 0.60], p = 0.216, see Fig 5b). The heterogeneity between studies was high (I^2^ = 88.02%, p < 0.001).

##### 3.2.3.3 Qualitative analysis MX vs. BCT

Six additional studies indicate that females with BC treated with BCT report higher body image than those in the MX group (33–37,72). However, Kraus et al. (26) found greater body image satisfaction in the MX group, while two other studies did not report a significant difference between the BCT and MX groups in either a positive (38) or negative (39) direction.

### 3.3 Risk of bias

The risk of bias was assessed using NOS and ROBINS-I (see Tables 1-3). Out of the twenty-five studies assessed by NOS, four were determined to be high quality (8–10), sixteen were of moderate quality (5–7) and five were judged as low quality (0–4) based on the NOS criteria. Out of the fifteen studies assessed by ROBINS-I, twelve were determined to be of high quality, and three were of moderate quality based on the ROBINS-I criteria.

### 3.4 Publication bias

Publication bias was examined for all analyses of body image. The Egger’s tests were not significant for any of the analyses: females with BC vs. HC (k = 5, value = 1.781, p = 0.075), before vs during treatment (k = 13, value = 0.373, p = 0.709), MX vs BCT-negative (k = 6, value = 1.118, p = 0.263), and MX vs. BCT positive (k = 8, value =-0.471, p = 0.638) indicating no evidence for publication bias in these analyses.

## 4. DISCUSSION

In this study, we conducted a systematic review and meta-analysis to comprehensively examine the effects of BC and its treatments on body image. While breast cancer treatment is both life-saving and essential, it can have a significant impact on body image. Specifically, we addressed three key research questions: 1) How does the body image of females with breast cancer (BC) compare to that of healthy controls, 2) How does the body image of females with BC change before vs during treatment, and 3) How does body image differ between patients undergoing mastectomy (MX) and breast-conserving therapy (BCT), considering both negative and positive aspects?

### 4.1 Females with BC vs. Healthy controls

The meta-analysis examined body image differences between females with BC and healthy controls, revealing significant body image dissatisfaction among females with BC. Similarly, qualitative analysis of studies included in the systematic review demonstrated the same pattern. Hence, the literature consistently shows that females with BC experience higher levels of body image dissatisfaction, body image-related problems, and lower body acceptance compared to females without BC (10,18,24,25,27,28,49,50). This negative body image may be attributed to treatment-related changes such as weight gain, hair loss, breast loss, and scarring, all of which can adversely affect body image (27). Females with BC often perceive their bodies as a source of negative emotions, leading to lower self-esteem and difficulties in daily life (50), and these factors contribute to reduced body acceptance and more pronounced body image alterations. Both our findings and existing literature consistently show a strong link between poorer body image outcomes and the effects of cancer treatments, including chemotherapy, radiation therapy, endocrine therapy, and surgery. Meta-analysis is essential in uncovering deeper insights, as it not only consolidates evidence across studies but also helps address gaps and variability in the current research. This approach ensures a more thorough and reliable understanding of these treatment effects.

Nowadays, various types of treatments are chosen based on factors such as tumor size, cancer stage, and patient’s condition. These treatments have many common side effects but also unique ones. All of them affect body image, be it hair loss from chemotherapy, loss of fertility from tamoxifen therapy, or loss of a breast from mastectomy, which can negatively impact femininity, motherhood. Importantly, the impact on body image frequently extends well beyond the active treatment phase. Research indicates that many females with BC continue to experience persistent, or even increased body dissatisfaction during survivorship, as psychological adaptation often lags behind physical healing (73). Body image remains dynamic after treatment, influenced by enduring physical changes as well as emotional and social factors (74).

Psychosocial interventions have shown promise in supporting this adjustment, especially when aligned with individual characteristics and treatment experiences. A recent systematic review has shown that various interventions including cognitive behavioral therapy, mindfulness-based programs, self-compassion writing exercises, and body image–focused group therapies, can effectively reduce negative body image and promote positive aspects such as body appreciation and acceptance (75). For instance, programs like “Accepting your Body after Cancer” and guided imagery sessions have been shown to alleviate distress and enhance psychological resilience in females with BC (75,76). Tailoring these interventions to individual characteristics, cancer stage, and cultural background can further increase their relevance and impact. Embedding such strategies into clinical pathways from diagnosis through survivorship may help bridge the gap between research and practice, ultimately improving the psychological well-being and quality of life of females with BC.

Body image has psychosocial dimensions that are shaped by social norms and culture (77). Research indicates that social norms significantly influence cancer-related perceptions in females with BC, often leading to negative body image (78). Anagnostopoulos and Myrgianni (24) report that in Western cultures, women’s bodies are highly valued, with breasts playing a crucial role in both gender identity and overall body integrity. Our systematic review and meta-analysis included studies not only from Western countries but also from regions such as the Middle East and Latin America (see Figure 2). This highlights the global impact of breast cancer on body image, drawing on data from diverse cultural contexts rather than focusing solely on a Western perspective. Although subgroup analyses to directly compare cultural differences were not conducted, the inclusion of studies and participants from various countries provides valuable insights into potential variations across populations. This multicultural approach broadens the understanding of body image issues beyond the confines of Western culture, offering a more comprehensive and inclusive perspective in the field.

Emerging evidence suggests that culture not only contextualizes but also actively shapes how females experience body image following breast cancer. Yoo et al. highlight the role of sociocultural factors (such as education, economic background, and societal expectations) as key influences on body image distress in individuals with cancer (79). A recent systematic review further underscores that appearance-related norms, gender roles, and cultural values related to femininity can shape coping mechanisms and perceptions of bodily change (80). For instance, in collectivist societies, physical integrity may be closely tied to familial or marital roles, potentially intensifying body image concerns, whereas in individualistic societies, self-presentation and autonomy may be more strongly emphasized. These dynamics suggest that cultural context can act as a moderator in the relationship between cancer treatment and body image outcomes. Future meta-analyses should examine these differences systematically, using culturally sensitive measures and conceptual frameworks.

Puberty, pregnancy, and menopause are crucial hormonal transition phases in a female’s life, each marked by physical changes and symptoms that can profoundly affect body image (81). Importantly, BC may co-occur with all of these transition periods (82–84), adding complexity to simple comparisons of patients with healthy controls. Therefore, examining body image during these transitions and exploring the experiences of females with BC in these phases, is crucial to account for possible variability in body image perceptions among different groups of females with BC.

### 4.2 Treatment effects

The meta-analysis evaluated potential changes in body image before vs. during treatment, mainly including data from studies on chemotherapy (n=4), radiotherapy (n=3), and surgery (n=7). The analysis confirmed that before the start of treatment, females with BC had a significantly higher body image scores, i.e. more positive body image, than during treatment (26,30–32,48,52,58,59,85). In terms of different treatment modalities, studies have shown a significant decrease in body image in females with BC undergoing chemotherapy (86), radiotherapy (53), and post-surgery (54,58). Thus, independent of treatment modality, a decrease in body image was reported. These changes may be attributed to factors such as hair and breast loss, weight fluctuations, and scarring associated with the various treatment approaches (27).

Interestingly, El Haidari et al. (55) observed that married females with BC in Lebanon experienced a significant improvement in body image one day after breast-conserving surgery. Authors speculate that marital status may positively influence body image perceptions after surgery, possibly due to increased emotional resilience or supportive relationship dynamics within Lebanese culture (55).

A qualitative analysis of the studies included in the systematic review revealed varying findings: while some studies (30–32) supported the meta-analysis results, others found no significant differences in body image between the before and during treatment periods (57,60). This inconsistency may be explained by a shift in focus from physical appearance to survival concerns during treatment. Depending on patient characteristics, they may prioritize different factors affected by BC, such as physical appearance, loss of fertility, unhealthiness, or high responsibilities in their lives (57). Another possible reason is that in these females with BC, body image is not affected by the side effects of treatment, but rather by the impact of being diagnosed with BC (39,67). For this reason, changes in body image need to be assessed carefully and external factors should be taken into account.

### 4.3 Treatment modality (negative & positive direction)

MX and BCT are two primary treatment options for breast cancer (BC) (87). This meta-analysis investigated the differences in body image between females with BC who underwent MX and those who had BCT. It was hypothesized that the various effects of these two different treatment modalities on the breast, such as scarring or complete breast removal, affect body image differently. It was expected that MX would have a more negative effect on body image compared to BCT. The results indicate that MX is associated with significantly greater negative body image than BCT. However, in terms of the positive aspects of body image, no significant group differences were found. Recent literature presents inconsistent findings regarding body image following these two treatment options: while several studies indicate that females with BC who underwent MX report worse body image than those undergoing BCT (24,34–37,61,62,64,69,71,72,88), others showed no significant difference (38,39,65,67). Previous studies observed greater body stigma (89) and greater alteration to their body image (36,90) in the MX group. Studies have shown that the MX group faces challenges with clothing and overall appearance, as well as body shame due to the loss of their breast(s) (91,92), which may contribute to greater body concerns and alteration.

Conversely, Kraus (26) found that females with BC who underwent MX reported higher body image satisfaction compared to those who underwent BCT. The author suggested that, in life-threatening situations following a BC diagnosis, physical appearance may become less significant, and the MX group might experience a sense of relief from the cancer diagnosis, which could mitigate concerns about body image despite the loss of their breast(s) (26). While this interpretation provides valuable insights, it must be viewed with caution due to the small sample size in the study (MX = 15, BCT = 16) conducted in 1999. Furthermore, since the study was conducted over two decades ago, social perspectives, cultural attitudes, and advancements in treatment modalities may have significantly evolved, potentially influencing body image experiences differently in contemporary populations. This is particularly relevant considering that BCT techniques were not widely adopted until the mid-1990s, and the quality of these procedures has improved over time. It is also important to note that the studies evaluating BCT are not randomized, which could affect the reliability of the findings.

Furthermore, several studies suggest no significant differences in body image between patients undergoing MX or BCT (38,39,65,67). This may be due to individual differences and personal beliefs of females with BC, suggesting that body image concern may be influenced more by the cancer itself than by the type of treatment received (38,39,65,67). Furthermore, the lack of significant differences in body image outcomes between these two types of surgery underscores the importance of prioritizing body image concerns for all patients, regardless of the surgical approach (50,93).

Moreover, the mixed findings regarding the effects of BC and its treatment on body image highlight the need for further research that considers sociodemographic factors. Education level, marital status, age, treatment modalities, and patients’ own perspective on their illness may play an important role in their body image and perspective on cancer. Additionally, raising awareness about breast cancer, its treatment, and its potential impact on body image is essential to support patients in managing these challenges effectively. Providing psychoeducation to help patients manage potential side effects can improve their understanding of body image issues associated with changes in breast shape or loss after surgery and their ability to cope with BC (94,95).

This systematic review and meta-analyses are the first to offer a comprehensive multi-parameter quantitative synthesis of the impact of BC on body image. Our rigorous inclusion criteria and standardized review process, in accordance with following the PRISMA guidelines, contribute to the robustness and reliability of our findings.

### 4.4 Study Limitations

Nonetheless, several limitations should be considered when interpreting these results. First, the number of studies included in some analyses was relatively small, particularly in the comparison of body image between breast cancer patients and healthy controls (n=5). Second, the lack of differentiation between participants based on treatment duration (e.g., days, months, or years) is another limitation. Future studies should consider categorizing participants into multiple groups based on treatment duration to allow for a more nuanced understanding of the effects of treatment. Third, while most studies comparing females with BC to healthy controls collected data post-treatment, two studies included both pre-and post-treatment patients without distinguishing between these subgroups in their analyses or conclusions. This methodological limitation may have obscured potential differences in body image experiences across treatment stages. Additionally, the wide period of the studies, as well as advances in surgical techniques and therapies, may introduce variability in the results. Breast-conserving surgery became widespread in the mid-1990s, with high-quality procedures becoming standard only after 2000. Poor surgical quality before 2000 likely influenced body image outcomes, but because only one study before 2000 included both MX and BCT, sub analysis was not possible. Lastly, since surgical studies are not randomized, postoperative satisfaction may not be differentiated without any bias.

The considerable heterogeneity observed in all our meta-analyses can be attributed to several interrelated factors, including differences in patient characteristics, treatment protocols, and study methodologies across the included studies. Variability in age, disease stage, cultural background, and psychosocial factors likely influenced how body image was perceived and reported by participants. Additionally, treatment-related differences—such as variations in the type and timing of surgery, chemotherapy, and radiotherapy—may have further contributed to inconsistencies in body image outcomes. Methodological factors, including the use of diverse body image assessment tools (e.g., different questionnaires), study designs (e.g., cross-sectional or longitudinal), and sample sizes, may also play a significant role in driving heterogeneity. These variations might likely underlie the observed differences in body image outcomes among BC patients, resulting in significant heterogeneity across studies.

### 4.5 Clinical Implications and Future Direction

This systematic review and meta-analysis offer valuable insight into body image changes associated with BC and its treatment, providing a detailed scientific perspective on these effects.

Recognizing the complex effects of breast cancer and its treatment allows both patients and healthcare providers to anticipate the body image challenges that may arise throughout the BC treatment process. Providing healthcare providers with this insight is essential, as it enables them to support patients more effectively by integrating body image considerations into patient care. The findings can help identify the potential impact of BC and its treatments on body image, which underscores the need to incorporate body image considerations into psychoeducation and treatment plans, ultimately improving patient support and quality of life. By proactively addressing these concerns, healthcare providers can promote emotional resilience, support mental health, and improve the quality of life for females with BC, contributing to a more comprehensive care experience.

Future research should reduce heterogeneity by standardizing assessment tools, stratifying participants by key variables, and harmonizing protocols to enhance comparability and deepen understanding of the relationship between breast cancer, its treatments, and body image.

## 5. CONCLUSION

This meta-analysis offers valuable insight into body image changes associated with BC and its treatment, providing a detailed scientific perspective on these effects. The findings demonstrate that females with BC experience significantly poorer body image compared to females without BC. Additionally, BC patients report better body image before starting treatment than ongoing treatment phases, suggesting that treatment itself contributes to body image concerns. Furthermore, among females with BC, those who undergo MX tend to report worse body image outcomes compared to those receiving BCT, highlighting the differential impact of treatment types on body perception. Although BC treatment is essential, its impact on body image remains a significant challenge. This review highlights the need for comprehensive treatment that addresses these concerns to improve women’s overall well-being and quality of life.

## List of abbreviations

BC: Breast Cancer
BCT: Breast Conserving Therapy
CI: confidence intervals
HC: Healthy Controls MX Mastectomy
NOS: Newcastle-Ottawa scale
PRISMA: Preferred Reporting Items for Systematic Reviews and Meta-Analyses
PROSPERO: International Prospective Register of Systematic Reviews
RE: Random Effects
REML: Restricted Maximum Likelihood
ROBINS-I: Risk Of Bias in Non-randomized Studies of Interventions Scale

## Declarations

### Ethics approval and consent to participate

Not applicable.

### Consent for publication

Not applicable.

### CRediT authorship contribution statement

**Serenay Yazici Sarikaya:** Conceptualization, Data curation, Formal analysis, Investigation, Methodology, Software, Visualization, Writing – original draft, Writing – review & editing. **Rui Wang:** Conceptualization, Validation, Writing – review & editing. **Ann-Christin S. Kimmig:** Conceptualization, Validation, Writing – review & editing. **Sara Brucker:** Conceptualization, Validation, Writing – review & editing. **Markus Hahn:** Conceptualization, Validation, Writing – review & editing. **Anna Wikman:** Conceptualization, Methodology, Resources, Supervision, Validation, Writing – review & editing. **Birgit Derntl:** Conceptualization, Funding acquisition, Methodology, Project administration, Resources, Supervision, Validation, Writing – review & editing.

## Data Availability

The data supporting the findings of this study are available from the corresponding author upon
reasonable request.

## Acknowledgments

This work was performed as part of the International Research Training Group, “Women’s Mental Health Across the Reproductive Years” (IRTG 2804). The IRTG 2804 is funded by the German Research Foundation (DFG). The grant number is: GRK 2804/1. The authors would like to thank all those who provided additional information for this meta-analysis.

## Availability of data and materials

The data supporting the findings of this study are available from the corresponding author upon reasonable request.

